# Emergency Department Utilization After 60-Day Peripheral Nerve Stimulation Versus Usual Care for Refractory Occipital Neuralgia: A Prospective Propensity-Matched Cohort, Difference-in-Differences Analysis

**DOI:** 10.64898/2026.10.02.26364550

**Authors:** Cameron W. Harris, Eric T. Nguyen, Sergio Mosquera Limas, Samir J. Sheth, Robert Chow, Charles A. Odonkor

## Abstract

**Background:** Temporary 60-day percutaneous peripheral nerve stimulation (PNS) can relieve pain in refractory occipital neuralgia (ON), but whether it alters acute healthcare utilization relative to the natural trajectory of the disease is unknown, because prior studies have lacked a concurrent comparator.

**Methods:** We performed a secondary analysis of prospectively collected data from a tertiary interventional pain center. Adults with medically refractory ON (primary diagnosis occipital neuralgia, ICD-10-CM M54.81) who underwent 60-day occipital PNS (n = 42) were compared with concurrent patients who were clinically eligible for PNS but were denied insurance authorization and continued optimized non-PNS management (usual care; n = 63). Headache-related emergency department (ED) encounters—index (occipital neuralgia) and non-index (other headache/facial-pain diagnoses)—were ascertained over the 12 months before and after the treatment window via the electronic record and the statewide health information exchange. Patients were 1:1 propensity-matched on a priori confounders (age, sex, baseline total headache-related ED utilization, HIT-6, and baseline headache-medication use). The primary analysis was a difference-in-differences (DiD) comparison of the change in headache-related ED utilization, reported as the additive difference (Hodges– Lehmann estimate, distribution-free 95% CI) and, alongside, as a multiplicative rate ratio (RR).

**Results:** Thirty-six matched pairs were well balanced. Baseline headache-related ED utilization was nearly identical between matched groups (6.3 vs. 6.2 visits/year). Over the following year, PNS patients’ utilization fell to 2.5 while usual-care patients’ rose to 6.9. The DiD was 5.0 fewer headache-related ED visits per patient (95% CI 3.0–6.0; p < 0.001), corresponding to a rate ratio of 0.36 (95% CI 0.27–0.48; p < 0.001). Results were consistent in the full sample, in covariate-adjusted models, and after excluding controls denied for incomplete conservative-therapy trials. The effect was driven by non-index headache visits.

**Conclusions:** Relative to a concurrent, matched usual-care cohort whose headache-related ED utilization rose over time, 60-day occipital PNS was associated with a substantial reduction in acute-care use, strengthening the case for a role of PNS on healthcare utilization in refractory ON.

## 1. Introduction

Occipital neuralgia (ON) is a painful headache disorder characterized by recurrent paroxysmal, shooting, or stabbing pain involving the posterior scalp in the distribution of the greater, lesser, and/or third occipital nerves [1]. The development of ON is postulated to result from irritation of the occipital nerves through vascular, neurogenic, muscular/tendinous, and/or osteogenic mechanisms [2]. Although the epidemiology of ON remains incompletely characterized, a recent systematic review reported a frequency of 0.6% to 24.4% among adults with headache and facial pain presenting to tertiary care, with a predominance in females and a mean age of approximately 50 years [3].

Conservative management of ON includes physical therapy modalities and medications such as tricyclic antidepressants and antiepileptics. Interventional management can be considered when conservative measures are insufficient and includes local anesthetic injections, corticosteroid injections, botulinum toxin injections, pulsed radiofrequency, and neuromodulation for refractory disease [2]. For patients with refractory disease, occipital nerve stimulation (ONS) has emerged as a potential treatment option, although systematic reviews in 2015 and 2023 found only Class III evidence, underscoring the continued need for higher-quality investigation [4,5]. More recently, temporary percutaneous peripheral nerve stimulation (PNS) has provided a less invasive alternative: Salmasi et al. and Gutierrez et al. reported clinically meaningful improvements in pain and pain interference following 60-day occipital PNS, with benefits persisting after lead removal [6,7]. A case report similarly documented sustained relief following a 60-day temporary PNS course for refractory ON [8].

Healthcare utilization is an important but understudied dimension of headache burden. In a national analysis of more than 188 million ED encounters, over 6 million were headache-related, and nearly half involved computed tomography imaging [9,10]. Greater headache-related disability is independently associated with ED use [11,12], and recurrent ED presentation is itself a recognized high-burden phenotype, for which standardized definitions of frequent or “super-” utilization have been developed [13–15]. Yet whether PNS changes acute healthcare utilization in refractory ON—and, critically, how any such change compares with what would have happened *without* PNS—remains unknown. Prior reports, including single-arm analyses of this cohort, are uncontrolled: they can describe a reduction in ED visits following treatment but cannot separate a treatment effect from the natural trajectory of the disease, regression to the mean, or secular trends. Establishing a role for PNS on healthcare utilization therefore requires a concurrent comparator of comparable, refractory patients who did not receive PNS.

We addressed this gap using a natural experiment. Within a tertiary academic medical center, some patients clinically eligible for 60-day occipital PNS received it, while others were denied insurance authorization and continued optimized non-PNS (“usual care”) management over the same period. We compared the change in headache-related ED utilization between these groups using a propensity-matched, difference-in-differences (DiD) design, which estimates the treatment-associated change *over and above* the concurrent change in comparable untreated patients. We hypothesized that 60-day occipital PNS would be associated with a greater reduction in headache-related ED utilization than the trajectory observed under usual care.

## 2. Materials and Methods

### 2.1 Study Design and Cohorts

We performed a secondary analysis of prospectively collected data from a single tertiary interventional pain practice. The treatment cohort comprised 42 consecutive adults with medically refractory ON who underwent 60-day occipital PNS between 1 January 2023 and 30 January 2025 after obtaining insurance authorization. The comparator (“usual-care”) cohort comprised 63 concurrent adults who were clinically evaluated and considered eligible for 60-day occipital PNS during the same period but whose insurance authorization was denied, and who therefore continued optimized non-PNS management. Documented denial rationales were “experimental/investigational” classification (n = 33), conflation with other stimulation modalities (n = 17), a required trial-to-permanent pathway (n = 6), and incomplete exhaustion of conservative therapy (n = 7). All patients carried a primary diagnosis of occipital neuralgia (ICD-10-CM M54.81), were ≥18 years of age, and had refractory disease despite prior conservative management. Usual care consisted of combinations of oral medications, physical therapy, trigger-point injections, transcutaneous electrical nerve stimulation (TENS), and occipital nerve blocks, at the discretion of the treating clinician.

This design constitutes a natural experiment: because PNS eligibility was clinically established in both groups and the determinant of treatment was payer authorization rather than disease severity, the comparator approximates the counterfactual trajectory of comparable refractory ON patients who did not undergo PNS. The study is a within-person, pre-versus-post comparison in each group, contrasted between groups (difference-in-differences); it is observational and non-randomized, and residual confounding cannot be excluded.

### 2.2 Peripheral Nerve Stimulation

Treatment-cohort patients underwent temporary percutaneous PNS targeting the occipital nerve(s) for a planned 60-day period using the SPRINT PNS System (SPR Therapeutics, Cleveland, OH, USA), consistent with our prior work [16]. Percutaneous leads were placed targeting the greater and/or lesser occipital nerves under ultrasound or fluoroscopic guidance; laterality and lead number were determined by each patient’s symptom distribution. Stimulation used a charge-balanced biphasic waveform (1–30 mA, 10–200 µs, 5–150 Hz), titrated to comfort, delivered continuously for up to 60 days, after which leads were removed.

### 2.3 Healthcare Utilization and Outcome Definitions

Headache-related ED utilization was ascertained from the electronic medical record supplemented by the statewide health information exchange (Connecticut’s Health Information Exchange, CONNIE; https://www.conniect.org), which captures ED and urgent-care encounters across participating sites statewide; the CONNIE query spanned the full 12-month pre- and post-treatment windows for every patient. For PNS patients, the follow-up window began after device removal. Each presentation was counted as a single unique encounter, with same-day registrations consolidated. Using prespecified ICD-10-CM codes, *index headache-related ED visits* were encounters coded for occipital neuralgia (M54.81); *non-index headache-related ED visits* were encounters coded for other headache or facial-pain diagnoses (migraine, G43.\; *other headache syndromes, G44*.*\*, including cervicogenic headache G44.86, tension-type G44.2x, post-traumatic G44.3x, trigeminal autonomic cephalalgias G44.0x, hemicrania continua G44.51, new daily persistent headache G44.52, and other specified syndromes G44.89; unspecified headache, R51.9; trigeminal neuralgia, G50.0; and atypical facial pain, G50.1), excluding M54.81 and generic cervicalgia (M54.2). *Total headache-related ED visits*, the primary outcome, were the sum of index and non-index encounters. Headache-related disability was assessed with the Headache Impact Test-6 (HIT-6).

### 2.4 Propensity Matching

To construct comparable groups, patients were 1:1 matched on a propensity score estimated by logistic regression. Matching covariates were specified a priori as established confounders of ED utilization and of treatment access—age, sex, baseline total headache-related ED utilization, baseline HIT-6, and baseline headache-medication use—and were not selected on the basis of observed imbalance [17]. Baseline headache-related ED utilization was included because, in a difference-in-differences framework, comparability of the pre-period outcome level underwrites the parallel-trends assumption. Matching was performed by nearest-neighbor selection on the logit of the propensity score within a caliper of 0.2 standard deviations, with ties broken by Mahalanobis distance on the matching covariates, without replacement. Balance was assessed by standardized mean differences (SMDs) before and after matching, with |SMD| < 0.25 considered acceptable (Figure 2).

**Figure 1.**
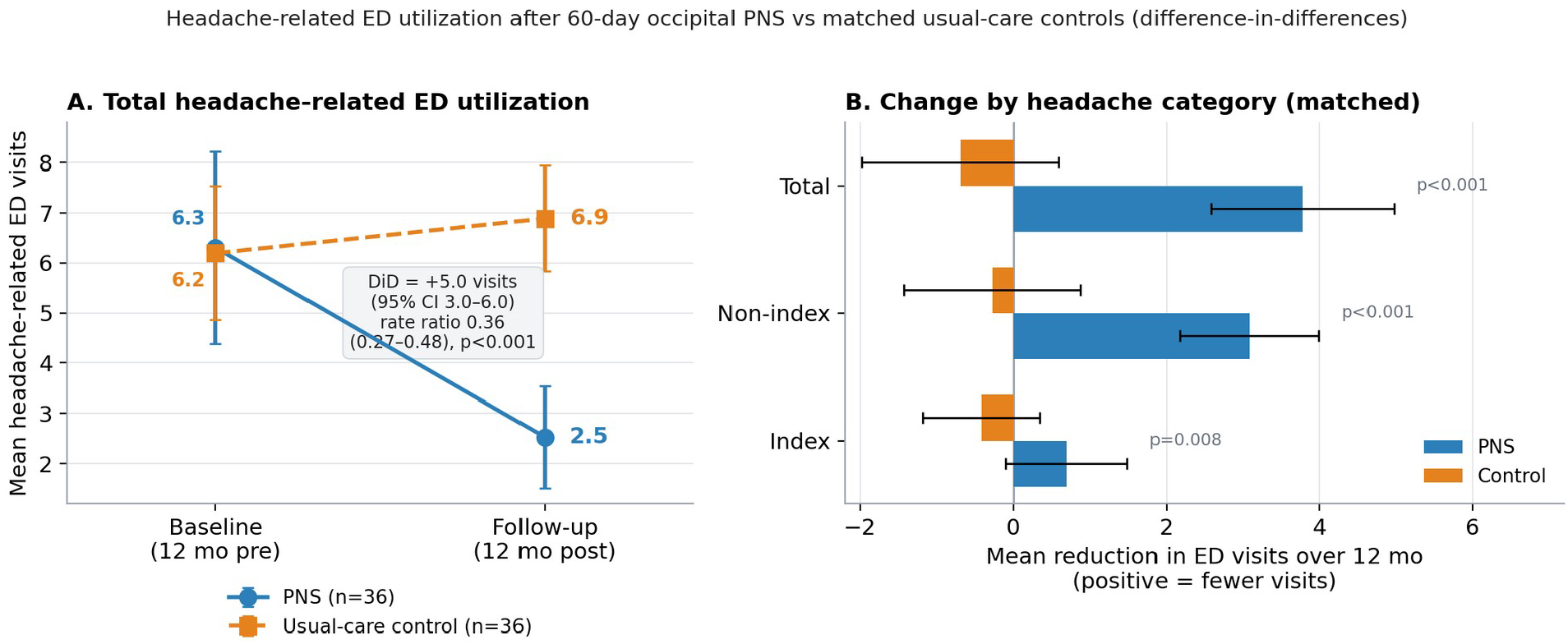
Difference-in-differences in headache-related ED utilization.(**A**) Mean total headache-related ED visits at baseline and 12-month follow-up in the matched cohort; the groups begin at an essentially identical level, after which PNS utilization falls and usual-care utilization rises. (**B**) Mean within-person reduction by headache category (positive = fewer visits), by group. Error bars are 95% confidence intervals.

**Figure 2.**
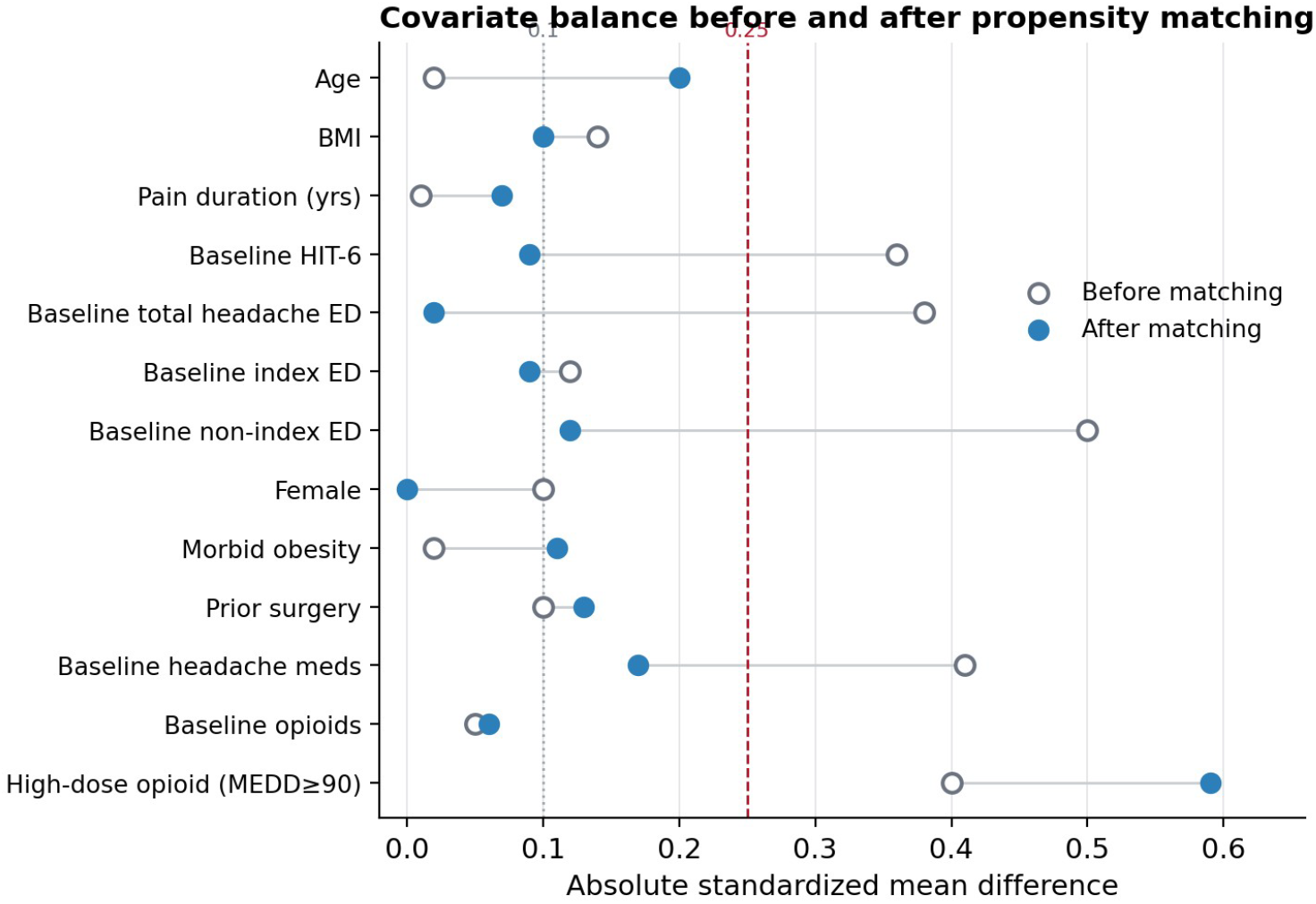
Covariate balance before and after propensity matching (Love plot). Absolute standardized mean differences for each matching and descriptive covariate. The dashed line at 0.25 marks the conventional threshold below which groups are considered well balanced (lower is better). Matching reduced imbalance below 0.25 for all covariates except high-dose opioid use, which was carried into adjusted analyses.

### 2.5 Statistical Analysis

The primary estimand was the difference-in-differences (DiD): the between-group difference in the within-person change (baseline minus follow-up) in headache-related ED utilization [18]. The DiD was reported, as the lead measure, on the additive scale in visits/year, using the Hodges–Lehmann estimator of the between-group difference in location with a distribution-free 95% CI and a two-sided Wilcoxon rank-sum (Mann– Whitney) test. Alongside the additive estimate, the DiD was also expressed on the multiplicative scale as a rate ratio (RR) from a generalized estimating equation (GEE) Poisson model with an exchangeable working correlation and cluster-robust (patient-level) standard errors, in which the treatment-by-period interaction term is the DiD; a rate ratio below 1 indicates a greater proportional reduction with PNS. Index and non-index components were analyzed identically. Within-group changes were tested with Wilcoxon signed-rank tests.

The primary analysis was performed in the matched cohort. Prespecified robustness analyses comprised (1) the full unmatched sample; (2) a covariate-adjusted additive DiD estimated by change-score regression with heteroskedasticity-robust (HC3) standard errors, adjusting for age, sex, baseline HIT-6, baseline headache-medication use, and high-dose opioid use (morphine-equivalent daily dose ≥90 mg), constituting a doubly robust analysis when combined with matching; and (3) a sensitivity analysis excluding usual-care patients denied for incomplete conservative-therapy trials, who might be less refractory. Given the sample size, no group-by-time model with extensive covariate sets was fitted, to avoid overfitting. Two-sided p < 0.05 defined significance. Analyses used Python (statsmodels, scipy).

## 3. Results

### 3.1 Cohorts and Balance

The treatment cohort (n = 42) and usual-care cohort (n = 63) were similar in age (63.4 vs. 63.1 years), sex (67% vs. 62% female), and pain duration (5.3 vs. 5.3 years). Before matching, the treatment cohort had higher baseline HIT-6 (50.8 vs. 47.4; SMD 0.36), higher baseline total headache-related ED utilization (8.0 vs. 5.5; SMD 0.38), lower headache-medication use (57% vs. 76%; SMD −0.41), and higher high-dose opioid use (29% vs. 13%; SMD 0.40) (Table 1). Propensity matching yielded 36 matched pairs; after matching, all covariates were balanced (|SMD| < 0.25) except high-dose opioid use, which remained higher in the PNS group (31% vs. 8%; SMD 0.59) and was carried into adjusted analyses (Figure 2, Table 1). Critically, baseline total headache-related ED utilization was nearly identical in the matched groups (6.3 vs. 6.2 visits/year), establishing a common pre-treatment trajectory for the DiD comparison.

**Table 1.** Baseline characteristics before and after propensity matching. Values are mean or %. SMD = standardized mean difference (treatment minus control).

| Characteristic | PNS (n = 42) | Control (n = 63) | SMD | PNS matched (n = 36) | Control matched (n = 36) | SMD |
| --- | --- | --- | --- | --- | --- | --- |
| Age (years) | 63.4 | 63.1 | 0.02 | 63.7 | 67.2 | −0.20 |
| Female (%) | 66.7 | 61.9 | 0.10 | 63.9 | 63.9 | 0.00 |
| BMI | 29.2 | 30.1 | −0.14 | 29.5 | 30.2 | −0.10 |
| Pain duration (years) | 5.31 | 5.33 | −0.01 | 5.33 | 5.56 | −0.07 |
| Morbid obesity (%) | 35.7 | 36.5 | −0.02 | 36.1 | 41.7 | −0.11 |
| Prior surgery (%) | 28.6 | 33.3 | −0.10 | 22.2 | 27.8 | −0.13 |
| Baseline headache-medication use (%) | 57.1 | 76.2 | −0.41 | 58.3 | 66.7 | −0.17 |
| Baseline opioid use (%) | 40.5 | 42.9 | −0.05 | 44.4 | 41.7 | 0.06 |
| High-dose opioid, MEDD ≥ 90 (%) | 28.6 | 12.7 | 0.40 | 30.6 | 8.3 | 0.59 |
| Baseline HIT-6 | 50.8 | 47.4 | 0.36 | 49.6 | 50.4 | −0.09 |
| Baseline total headache-related ED visits | 8.02 | 5.49 | 0.38 | 6.31 | 6.19 | 0.02 |
| — index (occipital neuralgia) | 2.40 | 1.95 | 0.12 | 1.53 | 1.83 | −0.09 |
| — non-index (other headache) | 5.62 | 3.54 | 0.50 | 4.78 | 4.36 | 0.12 |

### 3.2 The Natural Trajectory of Refractory ON Under Usual Care

Over the 12-month observation period, the two groups moved in opposite directions (Figure 1). In the full sample, usual-care patients’ headache-related ED utilization *increased* significantly, from 5.5 to 6.5 visits/year (mean change +0.98 visits; Wilcoxon p = 0.007), whereas PNS patients’ utilization fell from 8.0 to 3.5 (−4.57 visits; p < 0.001). The same divergence was seen in the matched cohort, in which the groups began at an essentially identical baseline: usual-care utilization rose from 6.2 to 6.9 visits/year while PNS utilization fell from 6.3 to 2.5 (Table 2, Figure 1). Thus, rather than remaining static, acute-care use in comparable refractory ON patients who did not receive PNS tended to worsen over time—an observation that provides the natural-history context against which the treatment effect must be judged.

**Table 2.**
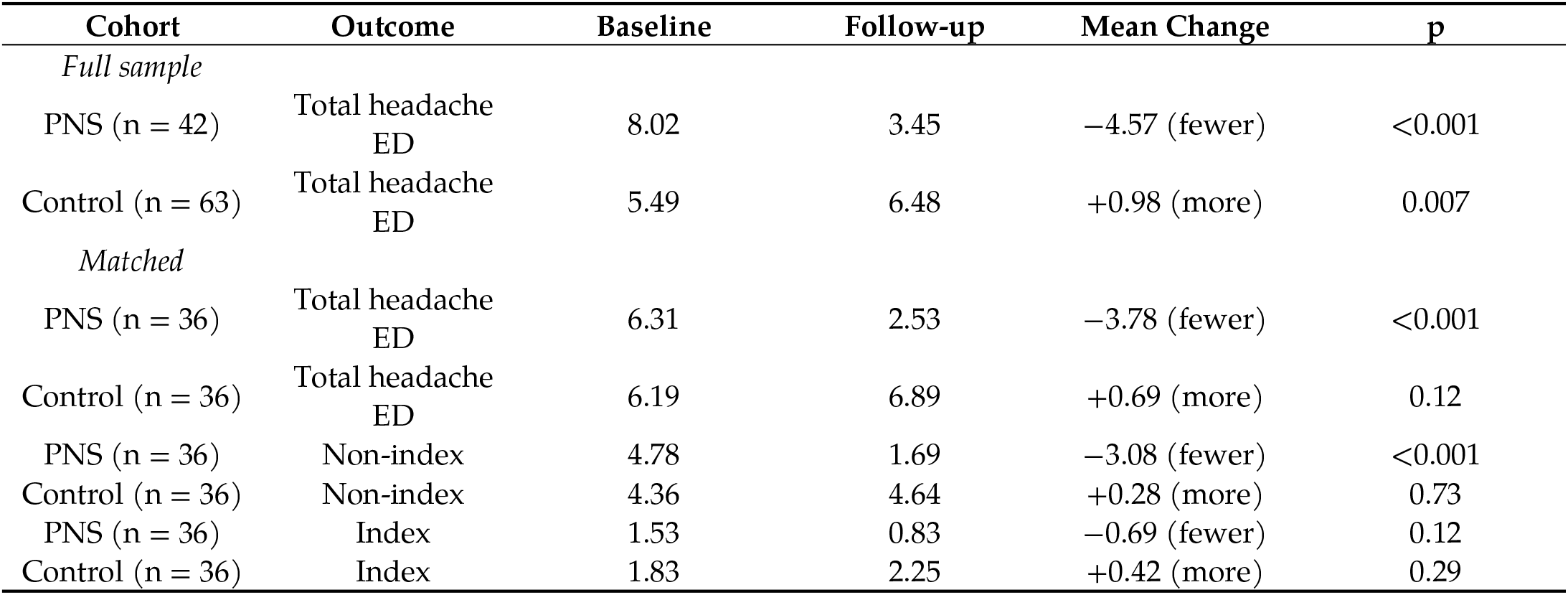
Within-group change in headache-related ED visits (baseline → 12-month follow-up). Positive reduction = fewer visits. *p*-values are two-sided Wilcoxon signed-rank.

| Cohort | Outcome | Baseline | Follow-up | Mean Change | *p* |
| --- | --- | --- | --- | --- | --- |
| *Full sample* |  |  |  |  |  |
| PNS (n = 42) | Total headache ED | 8.02 | 3.45 | −4.57 (fewer) | <0.001 |
| Control (n = 63) | Total headache ED | 5.49 | 6.48 | +0.98 (more) | 0.007 |
| *Matched* |  |  |  |  |  |
| PNS (n = 36) | Total headache ED | 6.31 | 2.53 | −3.78 (fewer) | <0.001 |
| Control (n = 36) | Total headache ED | 6.19 | 6.89 | +0.69 (more) | 0.12 |
| PNS (n = 36) | Non-index | 4.78 | 1.69 | −3.08 (fewer) | <0.001 |
| Control (n = 36) | Non-index | 4.36 | 4.64 | +0.28 (more) | 0.73 |
| PNS (n = 36) | Index | 1.53 | 0.83 | −0.69 (fewer) | 0.12 |
| Control (n = 36) | Index | 1.83 | 2.25 | +0.42 (more) | 0.29 |

### 3.3 Difference-in-Differences: Primary Analysis

In the matched cohort, 60-day occipital PNS was associated with a reduction in total headache-related ED utilization of 5.0 visits per patient per year beyond the concurrent usual-care trajectory (Hodges–Lehmann DiD 5.0 visits; 95% CI 3.0–6.0; p < 0.001). Expressed multiplicatively, the proportional change with PNS was 64% greater than under usual care (rate ratio 0.36; 95% CI 0.27–0.48; p < 0.001) (Table 3, Figure 1). The effect was carried by non-index (other-headache) ED encounters (additive DiD 3.0 visits, 95% CI 2.0–5.0; RR 0.33, 95% CI 0.23–0.48; both p < 0.001). For index (occipital neuralgia) encounters, which were infrequent, the additive DiD was not significant (0.0 visits, 95% CI 0.0–1.0; p = 0.18) while the multiplicative estimate was (RR 0.44, 95% CI 0.24–0.81; p = 0.008), consistent with a directionally concordant but count-limited signal.

**Table 3.** Difference-in-differences (PNS vs. usual care). Additive DiD = Hodges–Lehmann between-group difference in within-person change (visits/year), lead measure; rate ratio = multiplicative DiD (GEE Poisson treatment × period interaction, cluster-robust). RR < 1 favors PNS. For the index outcome, two *p*-values are shown (additive / multiplicative).

| Analysis / Outcome | Additive DiD, Visits (95% CI) | Rate Ratio (95% CI) | *p* |
| --- | --- | --- | --- |
| *Primary — matched (36* |  |  |  |
| *pairs)* |  |  |  |
| Total headache ED | 5.0 (3.0 to 6.0) | 0.36 (0.27 to 0.48) | <0.001 |
| — non-index | 3.0 (2.0 to 5.0) | 0.33 (0.23 to 0.48) | <0.001 |
| — index | 0.0 (0.0 to 1.0) | 0.44 (0.24 to 0.81) | 0.18 / 0.008 |
| *Full sample (42 vs. 63)* |  |  |  |
| Total headache ED | 5.0 (4.0 to 7.0) | 0.36 (0.29 to 0.46) | <0.001 |
| — non-index | 4.0 (3.0 to 5.0) | 0.33 (0.24 to 0.46) | <0.001 |
| — index | 0.0 (0.0 to 1.0) | 0.44 (0.27 to 0.72) | 0.11 / <0.001 |
| *Adjusted (full, change-score ANCOVA)* |  |  |  |
| Total headache ED | 5.6 (3.4 to 7.7) | — | <0.001 |
| *Matched, adjusted for MEDD ≥ 90* |  |  |  |
| Total headache ED | 4.2 (2.2 to 6.2) | — | <0.001 |
| *Sensitivity — exclude denial reason 4* |  |  |  |
| Total headache ED (full) | 5.0 (3.0 to 6.0) | 0.38 (0.30 to 0.49) | <0.001 |
| Total headache ED (matched) | 4.0 (2.0 to 5.0) | 0.39 (0.29 to 0.54) | <0.001 |

### 3.4 Robustness

Estimates were stable across specifications (Table 3). In the full unmatched sample, the total headache-related DiD was 5.0 visits (95% CI 4.0–7.0; RR 0.36, 95% CI 0.29–0.46; p < 0.001). A covariate-adjusted change-score model gave an additive DiD of 5.6 visits (95% CI 3.4–7.7; p < 0.001). Because high-dose opioid use remained imbalanced after matching, we confirmed robustness by adjusting the matched analysis for it (additive DiD 4.2 visits, 95% CI 2.2–6.2; p < 0.001); notably, high-dose-opioid patients had slightly *lower* baseline ED utilization, indicating that this residual imbalance, if anything, biased the estimate toward the null. Excluding the 7 usual-care patients denied for incomplete conservative-therapy trials did not materially change the result (full-sample DiD 5.0 visits, RR 0.38, 95% CI 0.30–0.49; matched DiD 4.0 visits, RR 0.39; both p < 0.001).

## 4. Discussion

In this propensity-matched, difference-in-differences analysis of patients with medically refractory ON, 60-day occipital PNS was associated with a reduction of approximately five headache-related ED visits per patient per year—about a two-thirds greater proportional reduction (rate ratio 0.36)—relative to a concurrent cohort of comparable patients who were eligible for PNS but continued usual care after insurance denial. The controlled design is the central advance over prior uncontrolled reports: by anchoring the treatment group’s change to the contemporaneous experience of similar untreated patients, it isolates the component of the reduction that is attributable to more than the passage of time.

### 4.1. The Natural Trajectory of Refractory ON Matters

The most instructive finding is what happened to the usual-care group. Far from remaining stable, their headache-related ED utilization rose over the year (significantly so in the full sample). Refractory ON that is not effectively treated appears to follow a worsening acute-care trajectory, plausibly reflecting disease persistence, escalating crises, and repeated reliance on emergency services in the absence of definitive relief. This has two consequences. First, it means that uncontrolled pre/post analyses—including our own earlier single-arm work—may *understate* rather than overstate the association, because they implicitly compare the treated group against an assumed-flat baseline rather than against the upward drift that actually occurs. Second, it reframes the benefit of PNS: the treatment is not merely lowering an elevated number but bending a rising curve downward, widening the gap between treated and untreated patients over time.

### 4.2. A Role for PNS on Healthcare Utilization

Against this trajectory, the magnitude and consistency of the PNS-associated reduction—stable across matched, full-sample, covariate-adjusted, and sensitivity analyses, and robust to the one residually imbalanced covariate—suggest that temporary occipital PNS may do more than relieve pain in the short term; it may reduce downstream acute-care utilization relative to the expected course of the disease. This is consistent with prior work showing decreased ED use after neuromodulation in other chronic-pain populations [19], and it extends the pain and disability benefits reported for 60-day occipital PNS [6,7] into the domain of health-system burden. The signal was carried by non-index (other-headache) encounters, with a concordant but count-limited effect on index occipital-neuralgia visits; this pattern is expected given that index encounters were infrequent, and it is compatible with a broad reduction in headache-driven acute presentations following effective occipital neuromodulation.

### 4.3. Implications for Value and Access

These findings also speak to the policy context that generated the comparator. The usual-care group exists because of insurance denial—most commonly on “experimental/investigational” grounds. Beyond the patient-reported pain and disability measures that typically anchor outcome assessment in pain medicine [20], healthcare utilization is an outcome that payers, health systems, and patients all recognize. The observation that denied patients went on to use *more* emergency care, while matched patients who received PNS used markedly less, is directly relevant to coverage decisions and to the value proposition of temporary PNS, and suggests that restricting access to PNS may shift, rather than avoid, costs—toward emergency care.

### 4.4. Limitations

This study has important limitations. It is observational and non-randomized; although treatment assignment was determined by insurance authorization rather than by clinician-judged severity, unmeasured confounding may persist, and the propensity model can balance only measured covariates. High-dose opioid use remained imbalanced after matching; we addressed this by adjustment and by noting its bias-toward-null direction, but residual effects cannot be excluded. The difference-in-differences estimate relies on the parallel-trends assumption, which cannot be fully verified with a single pre-period; matching on baseline utilization and the concordance of results across specifications provide partial reassurance. The sample is from a single academic medical center and a single state (though outside-system encounters were captured through CONNIE), the matched sample is modest (36 pairs), and diagnostic coding of headache subtypes is subject to misclassification. Index occipital-neuralgia encounters were infrequent, limiting power for that component. Finally, usual care was not protocolized and varied across patients. Prospective, ideally randomized, studies with standardized follow-up and broader resource-use measures are needed to confirm these findings.

## 5. Conclusions

Relative to a concurrent, propensity-matched cohort of comparable refractory ON patients whose headache-related ED utilization rose over time under usual care, 60-day occipital PNS was associated with a reduction of approximately five ED visits per patient per year (rate ratio 0.36). By situating the treatment effect against the natural, worsening trajectory of untreated refractory ON, these controlled data strengthen the evidence for a role of temporary occipital PNS in reducing acute healthcare utilization and support prospective, randomized, double-blinded controlled confirmation.

## Data Availability

All data produced in the present study are available upon reasonable request to the authors

## Supplementary Materials

The following supporting information can be downloaded at: [URL to be inserted by editorial office]. Table S1: STROBE checklist for cohort studies

## Author Contributions

Conceptualization, C.A.O., S.M.L. and S.J.S.; methodology, C.A.O., S.M.L. and S.J.S.; formal analysis, C.W.H., E.T.N., R.C., S.M.L., S.J.S. and C.A.O.; investigation and data curation, C.A.O., S.M.L. and S.J.S.; writing—original draft preparation, C.W.H., E.T.N., R.C., S.M.L., S.J.S. and C.A.O.; writing—review and editing, all authors; supervision, C.A.O. All authors have read and agreed to the published version of the manuscript.

## Funding

This research received no external funding

## Institutional Review Board Statement

The study was conducted in accordance with the Declaration of Helsinki and approved by the Yale University Human Research Protection Program Institutional Review Board (protocol #2000027238)

## Informed Consent Statement

All participants in the treatment cohort provided written informed consent at enrollment in the parent study; the comparator cohort comprised patients evaluated in the same practice whose data were analyzed under the same approved protocol

## Data Availability Statement

The data presented in this study are available from the corresponding author upon reasonable request; the data are not publicly available due to privacy and institutional restrictions

## Conflicts of Interest

C.A.O. and S.J.S. are consultants for SPR Pain Relief. S.J.S. consults for SI Bone, Medtronic, and Nalu outside this submitted work. C.A.O. consults for Boston Scientific and has received grant funding from Boston Scientific paid directly to Yale University, outside the submitted work; this relationship is unrelated to the current investigator-initiated study, which was independently conducted without industry sponsorship. All other authors declare no conflicts of interest related to neuromodulation or the submitted work. The funders had no role in the design of the study; in the collection, analyses, or interpretation of data; in the writing of the manuscript; or in the decision to publish the results

